# Metagenomic Analysis and Taxonomy of Bacteria Identified in Primary Endodontic Infections

**DOI:** 10.1101/2024.01.21.24301564

**Authors:** Alma Konjhodzic, Lajla Hasic Brankovic, Irmina Tahmiscija, Samra Korac, Aida Dzankovic

**Affiliations:** Department of Dental Pathology with Endodontics, Faculty of Dentistry with Dental Clinical Center, University of Sarajevo, Bosnia and Herzegovina

**Keywords:** Endodontic Microbiome, Endodontic Infections, Next-generation Sequencing

## Abstract

The study explores the polymicrobial nature of primary endodontic infections using Illumina Next Generation Sequencing. Samples involved in research have been collected from root canals of the patients suffering from pulp and periapical inflammations with no history of endodontic interventions on affected teeth. The study revealed prevalence of different bacterial phyla, classes, orders, and species. Further work will show potential correlations between individual microbiotas and clinical diagnosis.

## INTRODUCTION

The polymicrobial nature of primary endodontic infections has traditionally been a popular topic of research. Conventional microbiological methods were used to cultivate and isolate bacteria in order to classify them taxonomically. The taxonomy was performed to identify the microbiome members and provide a more detailed understanding of the relationship between symptoms and the microbes responsible for their manifestation.

The microbiological culture method commonly employs agar plates as a medium. However, when the greatest sample dilution is used, it results in an insufficient representation of the obtained sample. Aside from the disadvantages of dilution, the process of identifying microorganisms based on their phenotypic traits had additional significant drawbacks. As a result of their inherent ambiguity and the fact that some species may show phenotypically convergent or divergent behaviour, phenotypical traits are hard to assess. Moreover, culture methods are unable to extract non-viable, uncultivable, and, in certain situations, even fastidious or fragile bacteria (1). One of the main problems is replicating the right conditions for growth, such as the absence of vital nutrients for bacterial proliferation or interference with bacterial intercommunication systems. The inability to cultivate bacteria may also be attributed to the presence of other species that produce chemicals that hinder the growth of the target microbe. The technique is deficient in sensitivity, incurs high costs, and requires significant labour. Additionally, the cultivation and identification results might take a considerable amount of time, ranging from several days to weeks (2).

The analysis of pathogens in intracanals has progressed from traditional culture-based laboratory methods to molecular approaches and currently to microbiome-based, next-generation sequencing (NGS) culture-independent procedures (2). This procedure allows the identification of previously unknown human pathogens (3). The primary method for identifying bacteria is through PCR amplification and subsequent sequencing of the 16S ribosomal RNA (16S rRNA) gene (4).

Illumina NGS technology is a powerful tool for microbiological studies, particularly in the field of endodontics. NGS offers several key advantages over traditional microbiological methods. These advantages include increased sensitivity, more extensive analysis, and the ability to generate quantitative diversity and uniformity of data (5).

The Illumina NGS technology is very sensitive and can identify many different types of bacteria, even when they are present in very small amounts or at levels that are hard to detect (6). The presence of a particular bacterium can be important in intricate microbial communities, such as those found in root canals. Certain species of bacteria can significantly influence the overall ecology of the community, even in low concentrations.

Illumina NGS offers accurate quantitative data for every bacterial species. The amount of specific bacteria may influence the progression of the disease as well as the frequency and intensity of symptoms.

In addition, next-generation sequencing enables the detection and quantification of microorganisms that are currently unknown. These findings have the potential to extend current knowledge and make a substantial contribution to understanding the development and progression of pulpal and periapical diseases.

It is especially important to recognize the shifts in microbial flora, such as the transition from mostly aerobic to predominantly anaerobic. Each of these variables can influence the progression of the disease, the success of treatment, and recovery. NGS Illumina enables continuous monitoring of dynamic changes in the microbial population over a specific time frame.

Nowadays, the microbiome profiles associated with various endodontic infections lack comprehensive characterization (5). Data about microbiome dynamics obtained from our research may contribute to the development of more efficient protocols for disinfection and medication of the root canals, reducing the need for endodontic revisions (1)(6).

## Materials and Methods

### Patient Selection

The samples were collected from patients undergoing primary endodontic treatment at the Clinic for Dental Pathology with Endodontics of the Faculty of Dentistry, University of Sarajevo. The Institutional Ethics Committee approved this research, No. 02-3-4-19-4-2/22. Patients with a history of autoimmune and systemic diseases and antibiotic treatments 30 days prior to endodontic treatment were excluded (1). All patients gave written consent for research.

### Sample Collection

Prior to endodontic treatment, the affected tooth was isolated with a rubber dam, and the crown of the tooth was disinfected with a 3% sodium hypochlorite solution (1). The trepanation and extirpation were performed with sterile instruments following standard operating procedures (6). The paper point was placed in the canal for 60 seconds and then transferred into RNase-free Eppendorf tubes, which were sent for further procedures (1).

Out of the total sample, 50% of the samples were collected from the canal with vital pulp, while 25% of the samples were taken from the canal with necrotic pulp that did not show any evident radiological changes in the apex. The remaining portion of the sample was obtained from root canals displaying evident radiological signs of periapical disease.

### Isolation and sequencing of genetic material

The initial stage involved the isolation of bacterial DNA from the sample. The DNA was sequenced using a Qubit fluorometer, and sequencing libraries were prepared. Library preparation begins with the amplification of polymerase chain reaction (PCR) to amplify the region of interest, followed by purification using AMPure XP beads and the addition of adapters using Nextera XT indices (Illumina, San Diego, CA, USA) (6). The next step was the validation and quantification of the libraries obtained. The resulting libraries were combined and pipetted into a cartridge that assembles into a MiniSeq (Illumina) Next Generation Sequencer (NGS). The last step was bioinformatics data processing using FASTQ sequences and various bioinformatics software and tools.

## Results

Out of 40 endodontic interventions with sample collections, eight samples have been successfully sequenced to this point. A significant percentage of the collected samples (predominantly samples collected from vital cases) exhibited excessively high levels of human DNA concentration, impeding the sequencing process.

It is important to note that sequencing for some of the samples is still in progress. In line with this, to preserve research integrity and a bias-free environment, sequencing findings will not be matched with specific clinical cases or demographics of patients until all results are available.

Figures 1 through 4 display the findings of research conducted at the Phyla, Classes, and Species levels.

**Figure 1.**
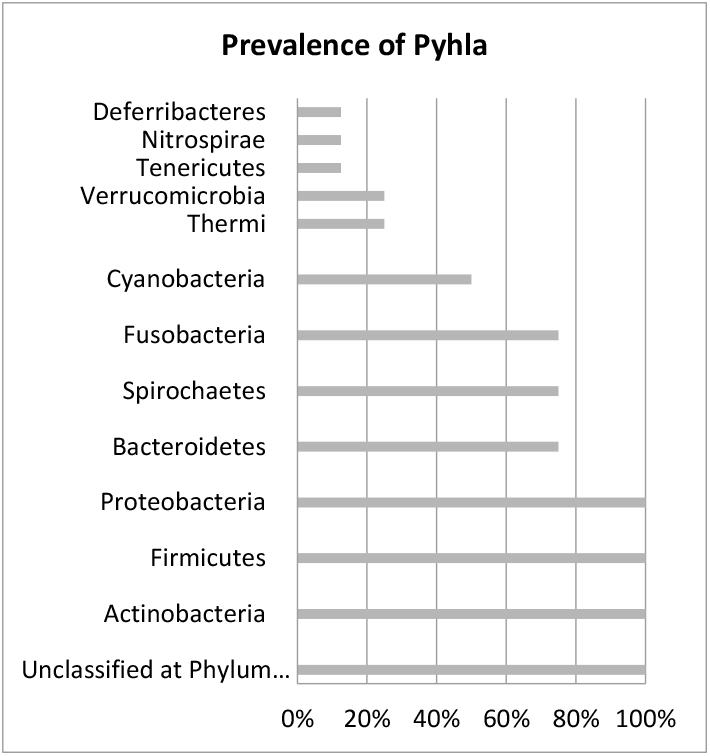
Prevalence of Phyla. The horizontal axis represents the percentage of samples containing members of a certain taxonomic level.

**Figure 2.**
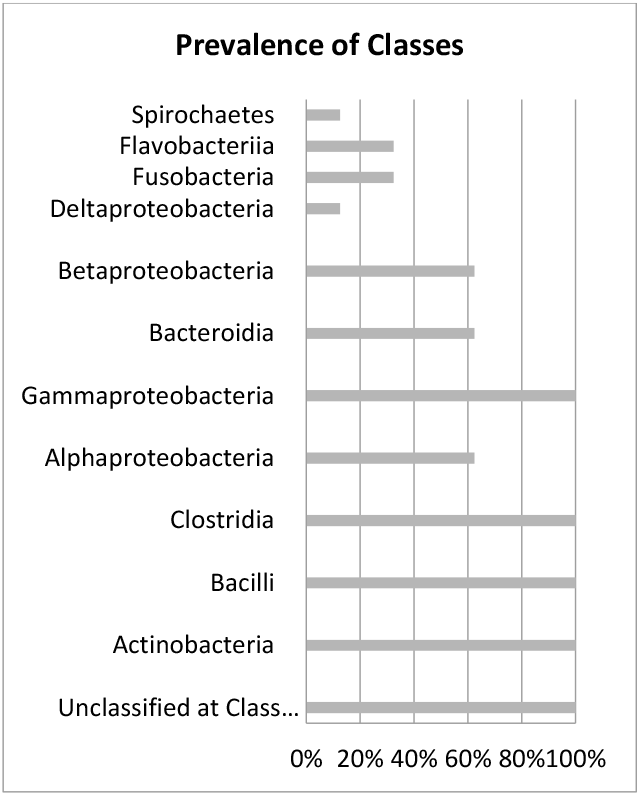
Prevalence of Classes.

**Figure 3.**
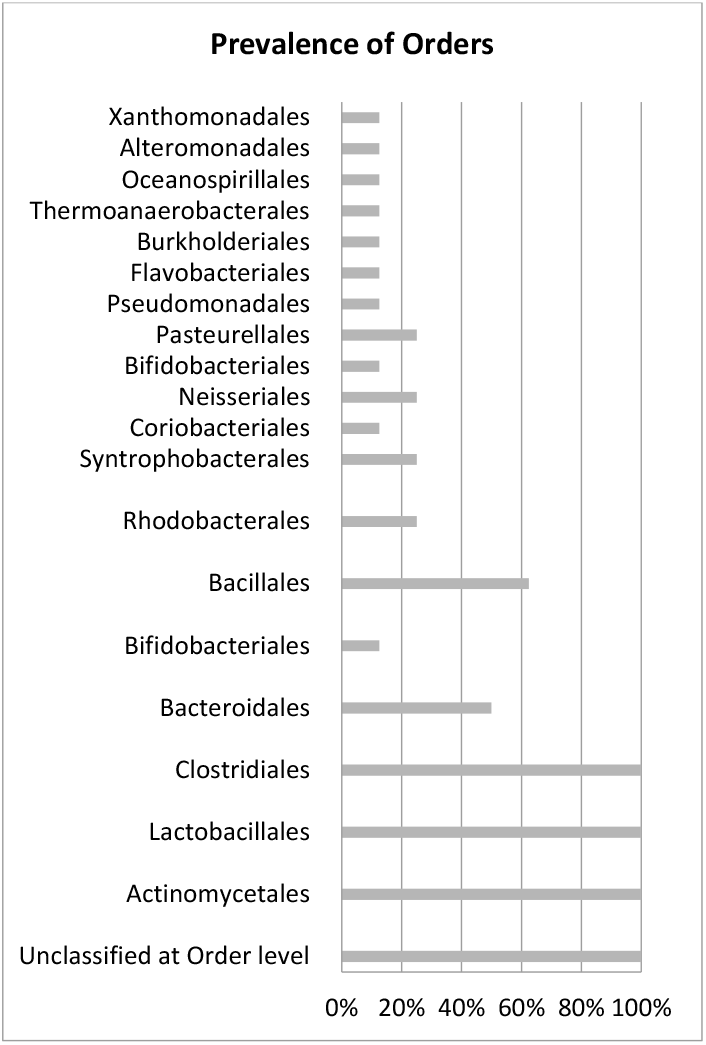
Prevalence of Orders.

**Figure 4.**
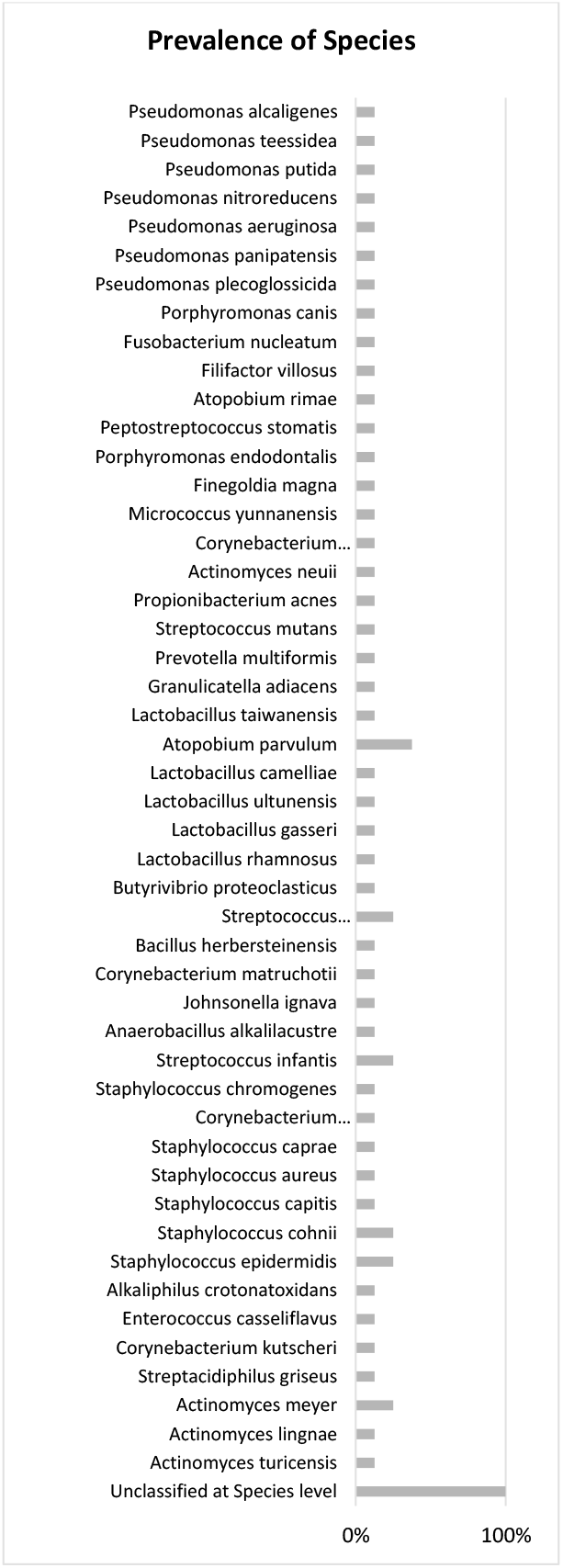
Prevalence of Species.

The phylum that exhibited the greatest abundance of bacteria in this study were Proteobacteria, Firmicutes, and Actinobacteria, followed by Bacteroidetes, Spirochaetes, and Fusobacteria (Figure 1).

### Prevalence of Pathogens

## Discussion

Forty samples of primary endodontic infections have been collected and sent for sequencing. At this moment, a total of 8 samples have been successfully sequenced, resulting in the data presented in the previous section. The phylum with the highest number of bacteria in this study were Proteobacteria, Firmicutes, and Actinobacteria, which corresponds to previous research findings (5)(6). The diversity of bacterial species in root canals can be attributed to the dynamic changes in the ecological environment that occur during the evolution of an infection, including the gradual depletion of oxygen and the necrosis of tissues (4). Due to the risk of compromising the free research environment, the individual results have not been cross-referenced with the characteristics of the patients. Considering this, further steps will be taken to find potential trends, identify probable trends, and create new treatment procedures based on these trends. Further discussion of the results, their quality, their potential impacts, and their relevance will be performed upon analysis of all samples at the end of this research project.

## Conclusion

At this phase of the research, it is not possible to draw any conclusions regarding the prevalence of certain pathogens in relation to clinical findings or demographics. The collective results of the research indicate that all forms of primary endodontic infections are associated with a wide range of microbiota. The final study’s conclusions will be confirmed upon its completion.

## Data Availability

All data produced in the present study are available upon reasonable request to the authors.

## Notes

### Competing Interest Statement

The authors have declared no competing interest.

### Funding Statement

This study was funded by Faculty of Dentistry University of Sarajevo, Ministry of Science, Higher Education and Youth of Canton Sarajevo

### Author Declarations

Ethics Committee of the Faculty of Dentistry of the University of Sarajevo gave ethical approval for this work. (Decision No. 02-3-4-19-4-2/22) Name of the Committee in official language of the institution: Eticki komitet Stomatoloskog fakulteta Univerziteta u Sarajevu

